# Advanced HIV disease among children and adolescents in HIV care in Uganda: prevalence, clinical outcomes, and rate of mortality

**DOI:** 10.1101/2025.11.24.25340918

**Authors:** Calvin Epidu, Rogers N. Ssebunya, Freddrick E. Makumbi, Edgar Sserunkuma, Emmanuel Tumwine, Patrick Kizza, Michael Juma, Henry Balwa, Betty Nsangi, Albert K. Maganda, Denise J. Birungi, Arthur G. Fitzmaurice, Dithan Kiragga

**Affiliations:** Baylor College of Medicine Children’s Foundation, Kampala, Uganda; School of Public Health Makerere University, Department of Epidemiology and Biostatistics, Kampala, Uganda; Division of Global HIV & TB, Global Health Center, US, Centers for Disease Control and Prevention, Kampala, Uganda

**Keywords:** HIV, AHD, Children, Adolescents

## Abstract

**Background:** People diagnosed with advanced HIV disease (AHD) are at high risk of increased mortality even after starting antiretroviral therapy (ART). We assessed AHD prevalence, clinical outcomes, and risk of mortality among children and adolescents living with HIV (CALHIV) in western Uganda.

**Methods:** We abstracted routinely-collected data of CALHIV aged 0-19 years from HIV clinic electronic medical records in 48 high-volume health facilities in two regions of western Uganda (Fort Portal and Hoima). Data for clients who initiated ART during January 2016—July 2023 were analysed. AHD was defined as a CD4 cell count <200 cells/μL, or WHO stage 3 or 4, or any child younger than five years of age living with HIV who had been on ART for more than 12 months and virally non-suppressed (≥1,000 copies). We used descriptive statistics (i.e., frequencies and percentages) to summarise prevalence and treatment outcomes. Kaplan-Meier curves were used to estimate overall survival and median time to death; log-rank tests were used to compare survival functions. A gamma-shared frailty model was used to determine factors associated with the rate of mortality. Effect measures were summarized using adjusted hazard ratios (aHRs) and their 95% confidence intervals (95%CI).

**Results:** A total of 5,143 CALHIV, including 3,067 (59.6%) females, with a median (interquartile range [IQR]) age of 10 (9) years were assessed. The overall prevalence of AHD was 18.2% (932/5,143) and varied by age—68.4% (0-4 years), 12.6% (5-9 years), 13.2% (10-14 years), and 7.7% (15-19 years). Just over half of the CALHIV diagnosed with AHD were active in care (51.5% [480/932]), about a quarter (26.4% [264/932]) had transferred out, 13.8% (129/932) were lost to follow-up, and 8.3% (77/932) had died. Survival was significantly higher in CALHIV who were not malnourished compared to those with malnutrition (p=0.001). Overall mortality rate among CALHIV with AHD was 3.41 (95% CI: 2.72-4.28) per 1,000 person-years and pronounced among those who had been on ART for three months or less (22.5; 95%CI: 17.0-29.8) compared to those above six months (0.8.2; 95%CI: 0.49 - 1.37).

**Conclusion:** Prevalence of AHD in CALHIV in western Uganda was within range compared to published adult-based studies. Risk of death differed by nutrition status and was high among those on ART three months or less. Early screening and management of malnutrition, as well as early ART initiation and adherence initiatives, might improve outcomes and reduce AHD-related mortality among CALHIV.

## Introduction

Amidst the global success in scaling up antiretroviral therapy (ART) coverage, children and adolescents living with HIV (CALHIV) remain hidden and left behind compared to adults. In 2023, nearly four in ten infants with HIV missed out on a timely diagnosis[1]. Each day in 2024, approximately 712 children aged 0-19 became infected with HIV and approximately 250 died from AIDS-related causes, mostly due to inadequate access to HIV prevention, care and treatment services[2]. The United Nations Children’s Fund (UNICEF) estimated close to 760,000—about 55% of the 1.38 million children aged 0-14 years living with HIV—were not on ART in 2024[3].

Treatment outcomes among children and adolescents 0-19 years old have improved since before the test-and-treat era but remain suboptimal. Retention-in-care rates, for example, improved globally after implementation of the 2017 guidelines to 55% in children aged 2–4 years, 65% in children aged 5–14 years, and 55% in children and adolescents aged 15–19 years[4]. Continuity in HIV care among CALHIV has also been suboptimal, with younger (<5 years) compared to older (5-9 years) pediatric patients being twice as likely to die within 90 days after ART initiation and older adolescents (15-19 years) demonstrating greater non-engagement in care in a large cohort in Tanzania[5]. Viral load suppression rates have also been shown to be lower compared to adults 18 years and older (i.e., 36% vs. 44% one year after ART initiation, 30% vs. 36% after year 2, and 24% vs. 29% after year 3)[6].

The World Health Organization (WHO) defines Advanced HIV Disease (AHD) as CD4 cell count <200 cells/mm^3^, WHO stage 3 or 4, or any child younger than five years of age living with HIV who had been on ART for more than 12 months and had an unsuppressed HIV viral load (>1,000 copies/ml)[7]. AHD is associated with poor treatment outcomes among PLHIV, including increased risk of opportunistic infections and death[8] as well as lower health-related quality of life (i.e., a worse physical or mental health status)[9]. According to the 2023 policy brief of the WHO[8], one fifth of the admitted AHD cases did not survive their hospital admission, and of those who survived, nearly a third died or were readmitted to the hospital within a year. “The path that ends AIDS: 2023 UNAIDS Global AIDS Update” report posited that AHD had become an increasing issue partly due to changing profiles of people with AHD including those who interrupt treatment and return to care[10]. The WHO recommends screening, treatment, and prophylaxis for major opportunistic infections; ART initiation within 7 days of diagnosis; and intensified adherence support as key interventions to reduce morbidity and mortality due to AHD.

Available evidence on the burden of AHD is mostly among adults, with a paucity of data on treatment outcomes including risk of mortality following diagnosis of AHD among CALHIV aged 0-19 years. Our study, therefore, set out to determine the prevalence, treatment outcomes, and rate of mortality following AHD diagnosis among CALHIV in mid-western Uganda.

## Materials and Methods

### Study design, setting, and population

This was a retrospective review of data of CALHIV (0-19 years) in HIV care in Hoima and Fort Portal regions in western Uganda. Data were abstracted from HIV clinic electronic medical records (EMRs) in 48 health facilities supported by the U.S. President’s Emergency Plan for AIDS Relief (PEPFAR) through the U.S. Centers for Disease Control and Prevention (CDC). Clients included in this evaluation were managed per Uganda’s HIV care and treatment guidelines[11]. All facilities included in this study provided the basic required HIV services including baseline CD4 count testing and screening and diagnosis of cryptococcal meningitis and tuberculosis (TB).

We included data for CALHIV (0-19 years) within the EMR who started ART during January 2016—July 2023 from 48 health facilities. These facilities with EMR contributed 80% of the active CALHIV during the April-June 2023 quarter. The analysis covers the test-and-treat period for all PLHIV operationalized in these regions beginning January 2016. Clients with missing date of ART initiation but with date confirmed HIV-positive were assigned the HIV-positive test date. We excluded CD4 count values evaluated before January 2016.

### Data collection procedures

With support from facility-based medical records assistants (MRAs), regional monitoring and evaluation officers, and health information system (HIS) officers, routinely-collected data were abstracted from EMRs in the health facilities within the two regions of Hoima and Fort Portal. Data were abstracted on 30 April 2024 and were fully anonymized before being made available to the research team. The key data variables abstracted included client sociodemographic characteristics, HIV diagnosis, baseline CD4 count, WHO clinical staging, TB status, cryptococcal meningitis status, and last clinic encounter date. The abstracted data covered five clinic visits. Data quality checks were conducted, and missing data were cross-referenced from registers and cleaned accordingly. Data were cleaned using Microsoft Excel and exported to STATA statistical software version 18.0 for analysis.

### Variable measurements

The main outcome of interest was AHD, defined as CD4 cell count <200cells/mm3 or WHO stage 3 or 4 in adults and adolescents, and all children under five years of age regardless of CD4 count[11]. However, children younger than five years who had been on ART for more than one year and were virally suppressed were not classified as AHD. Secondary outcomes of interest included active in care, died, transferred out (TO), and lost to follow-up (LTFU). CALHIV who reached 28 days after their scheduled clinic appointment date without returning to care were considered LTFU. All TO clients had a documented outcome as “Transferred out” in the EMR by the time of data abstraction. CALHIV were categorised as active if their last clinic encounter was within 28 days or their next clinic visit date was ahead of the date of data abstraction. Time-to-death was estimated from time of ART initiation to documented date of death, while administrative censoring was considered at month 60 if no event occurred before the 60-month follow-up. Other variables of interest included nutrition status and any confirmed stage 3 or 4 condition (tuberculosis and cryptococcal meningitis) or a classification of WHO clinical stage 3 or 4 at any of the last five clinic visits. The last clinic visit date was considered as date of transfer for clients documented as TO but with missing date. Nutrition status was based on mid-upper arm circumference (MUAC) measurements routinely done for CALHIV in HIV clinics i.e. color code “Red” for Severe acute malnutrition, “Yellow” for moderate acute malnutrition and “Green” for No malnutrition. For CALHIV with missing color codes, we used their weight for age z-scores to categorise their nutrition status.

### Statistical analysis

We conducted an exploratory analysis with descriptive statistics on all key variables of interest. AHD prevalence was defined as the number of clients classified as AHD divided by total number of CALHIV. Chi-square tests were used to assess associations between prevalence and clients’ sociodemographic and health facility characteristics. The rate of death was determined as the number of CALHIV who died during the study period divided by person-time-at-risk (pya) and reported per 1,000 pya. Kaplan-Meier method estimation was used to obtain the survival probabilities. Kaplan-Meier curves were used to estimate overall survival and median time to death for different levels of AHD status and other characteristics and the significance of the differences was assessed using log-rank tests. P-values of < 0.05 were considered significant. To determine independent factors associated with time-to-death, we fitted a proportional hazard model with a gamma shared frailty. Effect measures were summarized using adjusted hazard ratios (aHRs) and their 95% confidence intervals (95%CI).

### Ethics

This activity was reviewed by U.S. Centers for Disease Control and Prevention (CDC), and was determined to be non-research, and was conducted consistent with applicable federal law and CDC policy, 45 C.F.R. part 46, 21 C.F.R. part 56; 42 U.S.C. §241(d); 5 U.S.C. §552a; 44 U.S.C. §3501 et seq. An exemption from an Institutional Review Board (IRB) was granted by the Makerere University School of Public Health Higher Degrees, Research and Ethics Committee in Uganda. Consent for study participants was not required in this study since this was based on routinely captured secondary data in health facilities.

## Results

### Baseline individual characteristics

Table 1 shows both sociodemographic and clinical characteristics of study participants. High proportions of CALHIV who received treatment from lower-level health facilities (Health Centre III (HCIII), 50.2%), were categorised as WHO clinical stage 1 or 2 (93.7%), and were initiated on either non-nucleoside reverse transcriptase inhibitors (NNRTIs) (41%) or integrase inhibitors (37.1%) as their baseline ART regimens. Of those with documented viral load values, most (81.7%, 2,741/3,354) were virally suppressed. The proportion of CALHIV with history of TB diagnosis in this study population was 2.3% (117/5,143).

**Table 1.**
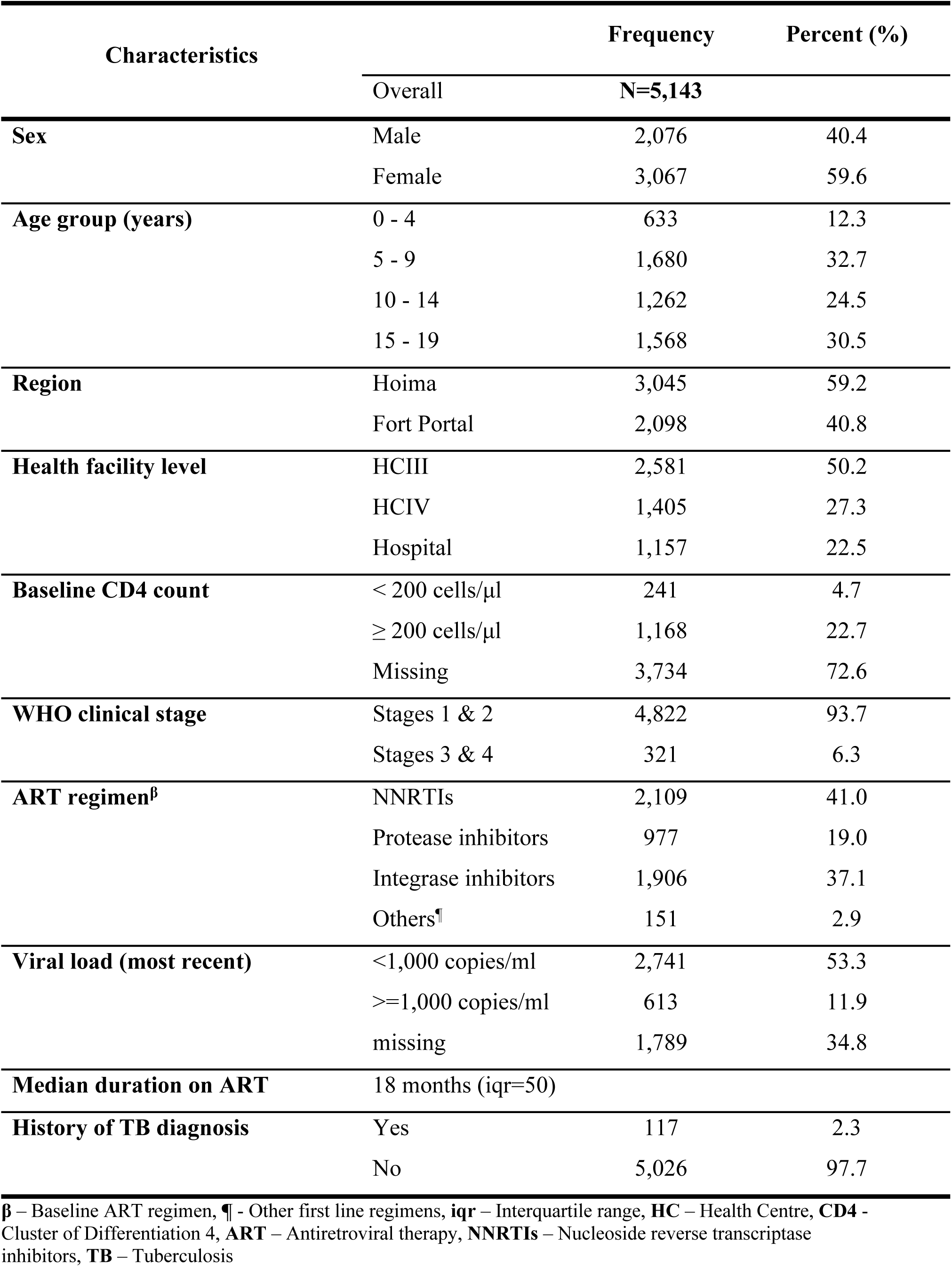
Characteristics of CALHIV assessed for AHD between January 2016 - July 2023 in two regions of Uganda.

### Prevalence of Advanced HIV Disease and treatment outcomes by individual characteristics

Table 2 highlights the prevalence of AHD, overall and stratified by client characteristics. Overall, the prevalence of AHD was 18.1% [17.1-19.2] (932/5,143) and highest among children 0-4 years (68.4%) [64.6-72.0]. The prevalence of AHD did not differ by region (Hoima at 18.5% (563/3,045) and Fort Portal at 17.6% (369/2,098)).

**Table 2.** Prevalence of Advanced HIV Disease (AHD) among CALHIV assessed between January 2016 - July 2023 in Hoima and Fortportal regions, Uganda.

|  |  | AHD (n/N) | AHD (%) | p-value |
| --- | --- | --- | --- | --- |
| **Characteristics** | Overall | 932/5,143 | 18.2 |  |
| **Sex** | Female | 510/3,067 | 16.6 | 0.001 |
|  | Male | 422/2,076 | 20.3 |  |
| **Age group (years)** | 0 - 4 | 433/633 | 68.4 | 0.001 |
|  | 5 - 9 | 212/1,468 | 12.6 |  |
|  | 10 - 14 | 167/1,262 | 13.2 |  |
|  | 15 - 19 | 120/1,568 | 7.7 |  |
| **Region** | Hoima | 563/3,045 | 18.5 | 0.205 |
|  | Fort portal | 369/2,098 | 17.6 |  |
| **Duration on ART<sup>p</sup>** | 0 - 6 | 383/1,582 | 24.2 | 0.001 |
|  | 7 - 12 | 189/642 | 29.4 |  |
|  | 13 - 24 | 90/630 | 14.3 |  |
|  | >24 | 270/2,289 | 11.8 |  |
| **Nutrition status** | No Malnutrition | 267/1,620 | 16.5 | 0.001 |
|  | Moderate Acute Malnutrition | 50/204 | 24.5 |  |
|  | Severe Acute Malnutrition | 175/601 | 29.1 |  |
|  | Missing | 440/2,718 | 16.2 |  |
| **ART regimen<sup>a</sup>** | NNRTIs | 113/677 | 16.7 | 0.033 |
|  | Protease Inhibitors | 112/499 | 22.4 |  |
|  | Integrase Inhibitors | 698/3,896 | 17.9 |  |
|  | Others<sup>¶</sup> | 9/71 | 12.7 |  |

<sup>p</sup> – Duration on months, <sup>a</sup> – Most recent clinic visit regimen, **ART** – Antiretroviral therapy, **NNRTIs** – Nucleoside reverse transcriptase inhibitors, <sup>¶</sup> - Second-line containing both integrase and protease inhibitors

More than half (61.4%) with AHD had been on ART for 12 months or less. Of those with data on nutrition status, 24.5% (50/204) and 29.1% (175/601) of CALHIV with moderate and severe acute malnutrition, respectively, had AHD.

Table 3 highlights HIV treatment outcomes among those with AHD. Overall, half (51.5%, 480/932) were active in care, 8.3% (77/932) had died, 13.8% (129/932) were LTFU, and 26.4% (246/932) had been TO from their primary HIV care facilities. The proportion of CALHIV who died varied by age group (i.e., 8.3% (36/433) among 0-4 years, 10.4% (22/212) among 5-9 years, 6.0% (10/167) among 10-14 years, and 7.5% (9/120) among those aged 15-19 years). The proportion of CALHIV with AHD who died were higher in Hoima Region (9.4%) compared to Fort Portal Region (6.5%) but this was not statistically significantly different (p=0.115. Among CALHIV who had been on ART for 6 months or less, a significantly higher proportion (15.9%, 61/383) died compared to those who had been on ART much longer (i.e. 4.8% (7-12 months), 2.2% (13-24 months) and 1.9% (>24 months)). A similarly high proportion (20.6%, 36/175) of CALHIV with AHD who had died also had severe acute malnutrition.

**Table 3:** Treatment outcomes among CALHIV diagnosed with Advanced HIV Disease between January 2016 - July 2023in Uganda.

| Variable |  | Total (N) | Active (n, %) | Died (n, %) | LTFU (n, %) | Transferred out (n, %) |
| --- | --- | --- | --- | --- | --- | --- |
| **Overall** |  | 932 | 480 (51.5) | 77 (8.3) | 129 (13.8) | 246 (26.4) |
| Sex | Male | 422 | 217 (51.4) | 43 (10.2) | 60 (14.2) | 102 (24.2) |
|  | Female | 510 | 263 (51.6) | 34 (6.7) | 69 (13.5) | 144 (28.2) |
| **Age group (years)** | 0 - 4 | 433 | 250 (57.7) | 36 (8.3) | 47 (10.9) | 100 (23.1) |
|  | 5 - 9 | 212 | 100 (47.2) | 22 (10.4) | 34 (16.0) | 56 (26.4) |
|  | 10 - 14 | 167 | 74 (44.3) | 10 (6.0) | 26 (15.6) | 57 (34.1) |
|  | 15 - 19 | 120 | 56 (46.7) | 9 (7.5) | 22 (18.3) | 33 (27.5) |
| **Region** | Hoima | 563 | 293 (52.0) | 53 (9.4) | 81 (14.4) | 136 (24.2) |
|  | Fort Portal | 369 | 187 (50.7) | 24 (6.5) | 48 (13.0) | 110 (29.8) |
| **Duration on ART<sup>p</sup>** | 0 - 6 | 383 | 107 (27.9) | 61 (15.9) | 68 (17.8) | 147 (38.4) |
|  | 7 - 12 | 189 | 124 (65.6) | 9 (4.8) | 18 (9.5) | 38 (20.1) |
|  | 13 - 24 | 90 | 51 (56.7) | 2 (2.2) | 13 (14.4) | 24 (26.7) |
|  | >24 | 270 | 198 (73.3) | 5 (1.9) | 30 (11.1) | 37 (13.7) |
| **Nutrition status** | No Malnutrition | 267 | 178 (66.7) | 8 (3.0) | 29 (10.9) | 52 (19.5) |
|  | Moderate Acute Malnutrition | 50 | 31 (62.0) | 2 (4.0) | 4 (8.0) | 13 (26.0) |
|  | Severe Acute Malnutrition | 175 | 55 (31.4) | 36 (20.6) | 38 (21.7) | 46 (26.3) |
|  | Missing | 440 | 216 (49.1) | 31 (7.1) | 58 (13.2) | 135 (30.7) |

<sup>p</sup> – Duration on months, **ART** – Antiretroviral therapy, **LTFU** – Lost to follow-up

### Survival experiences by age group and nutrition status among CALHIV with AHD

Figure 1 is the KM curve highlighting the probability of survival of CALHIV with AHD. The overall probability of survival was above 75% and varied by age group and nutrition status. Children 0-4 years old and 5-9 years old had lower survival experience compared to those 10 years and older, although this was not statistically significant (p-value=0.345). CALHIV with severe or moderate acute malnutrition had significantly lower survival compared to those without malnutrition (p-value=0.001). Generally, irrespective of age and nutritional status, death occurrences were highest within the first 15 months of starting ART and then plateaued. Deaths occurred even after 24 months of starting ART across all age groups and among those with malnutrition.

**Figure 1.**
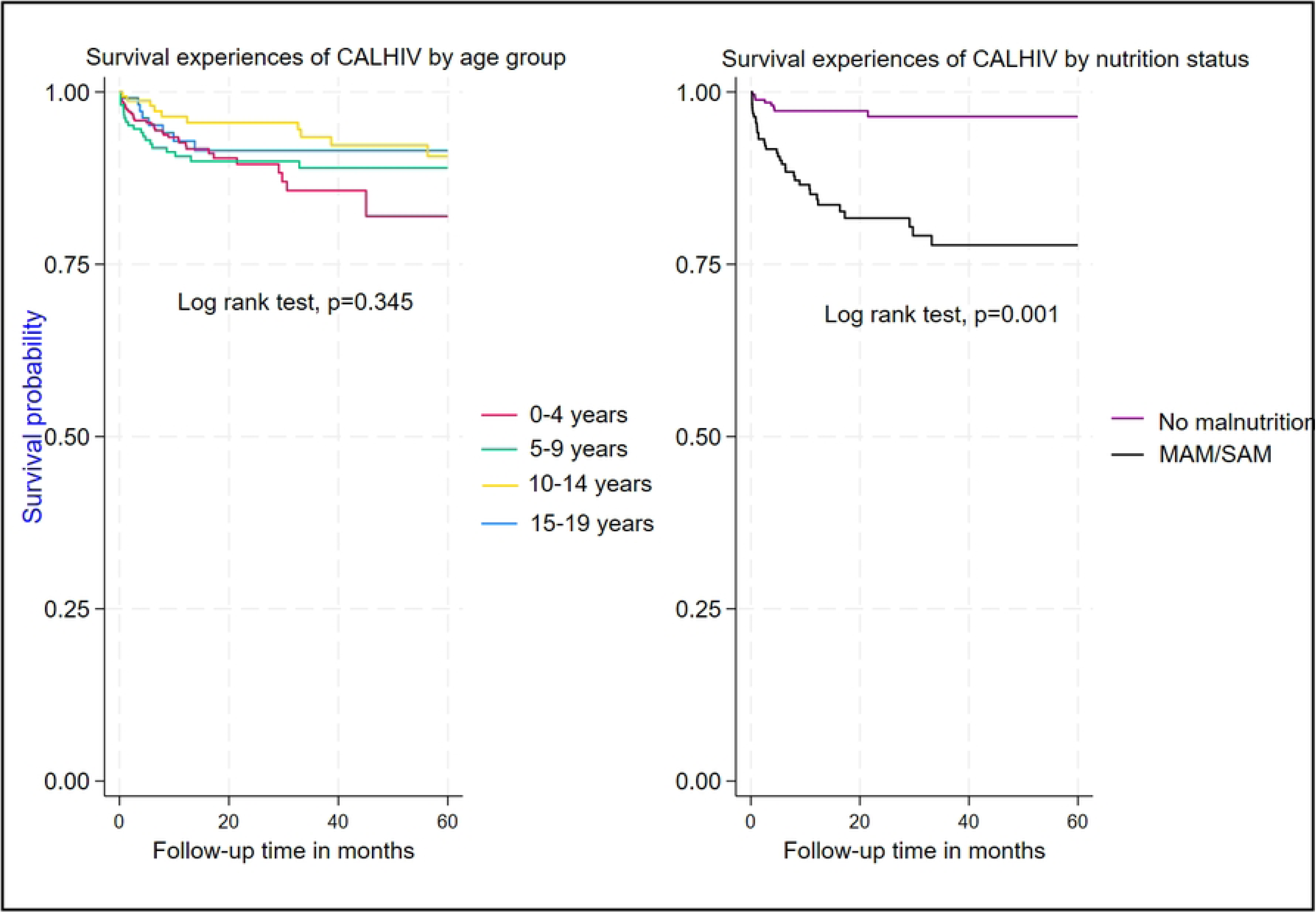
Survival by age group and nutrition status among CALHIV diagnosed with Advanced HIV Disease between January 2016 - July 2023 in Uganda.

### Mortality rate and associated factors of mortality among CALHIV with AHD

The overall mortality rate among CALHIV with AHD as shown in table4 was 3.41 (95%CI: 2.72-4.28) per 1,000 pya and predominantly seen among those who had been on ART for 3 months or shorter (22.5 [95%CI: 17.0-29.8] per 1,000 pya,), followed by CALHIV who had spent 4-6 months on ART (8.02 [95%CI: 4.44-14.49] per 1,000 pya). Males had a higher mortality rate compared to females (4.30 [95%CI: 3.18-5.82] vs. 2.68 [95%CI: 1.90-3.79] per 1,000 pya). The rate of death in children 0-4 years old was 5.33 (95%CI: 3.84-7.38) per 1,000 pya, 3.24 (95%CI: 2.09-5.03) per 1,000 pya among 5–9-year-olds, and 2.42 (95%CI: 1.21-4.84) per 1,000 pya among 15–19-year-olds. CALHIV with moderate and severe acute malnutrition were more likely to die (adj.HR=8.78; 95%CI: 2.72-28.34) compared to those who were not malnourished. CALHIV who had been on ART for <=3 months were also more likely to die compared to those who had been on ART for 6 months and longer (adj.HR=205.2; 95%CI: 43.1-977.3).

## Discussion

Our study showed that the overall prevalence of AHD among CALHIV was 18.2% and varied by clients’ characteristics, with children under five years old, those who had been on ART for ≤6 months, and those with malnutrition having high proportions. Overall mortality risk among those with AHD was 3.41 per 1,000 persons-years and higher among those with moderate and severe acute malnutrition. Approximately 13.8% and 26.4% among those with AHD had interrupted their treatment and TO, respectively. These results suggest that early screening and diagnosis of AHD and prompt ART initiation might reduce deaths. Particular care for CALHIV with malnutrition might not only address cause of mortality but also improve health outcomes among CALHIV with AHD overall.

The overall prevalence of AHD of 18.2% among CALHIV is within range compared to what has been reported elsewhere [12,13] among those at least 15 years olds but higher than in other studies[14] and unacceptable in this era of test-and-treat for all PLHIV. This finding suggests prophylactic, diagnostic, and therapeutic challenges among CALHIV, revealing potential targets for innovations to capture such clients early and to scale prophylaxis interventions for known opportunistic infections. Scale-up of TB preventive treatment (TPT) with isoniazid has faced implementation challenges especially with inadequate stock, and cryptococcal meningitis prophylaxis has been associated with inadequate health care provider knowledge and clinical competence to prescribe the recommended drugs. Additionally, skills to diagnose paediatric TB and cryptococcal meningitis are not readily available in most of the ART clinics especially lower level and rural facilities.

This is among the few studies that have explored the prevalence of AHD among children under 15 years old. Evidence documented in other published studies was among older adolescents (at least 15 years of age) and never included children and younger adolescents.

Our study also found that the majority (81%) of cases with AHD were among CALHIV nine years and younger. This is not surprising, as a number of children under five years old were considered to have AHD if they were on ART for less than12 months. On the other hand, our study had a number of CALHIV on optimised ART regimens (integrase inhibitors, i.e., DTG-based regimens)[15] and with CD4 counts of 200 cells/μL and above. This poses the question of whether to rethink the definition of AHD that includes all those under five-year-olds who have not been on ART for 12 months. That said, this slightly higher proportion of AHD among the younger age group could also point to HIV/AIDS-related stigma among parents or caregivers of such children in the community.

AHD prevalence was greater among males than females. There is evidence elsewhere[16] around sex differences in HIV disease progression and treatment outcomes, highlighting greater proportions of undetectable viral loads and survival among females compared to males. The current finding elicits a future research question of whether sex differentials, including potential effects of stigma, in communities influence male CALHIV.

The high proportion of CALHIV with AHD among those who had been on ART for 12 months or less is not surprising since this is the time when critical assessments are done including baseline CD4 count and TB screening. It is also within this critical period that CALHIV are adapting to the new reality of daily ART swallowing amidst existing HIV-related stigma. However, it is important to note that the proportion with AHD after two years on ART was surprising, because we would expect most of these clients to have stabilised with improved CD4 counts and no stage 3 or 4 AIDS-defining illnesses.

Our study also highlights that HIV treatment outcomes among CALHIV with AHD are not as good as expected, with about 22.1% either died or LTFU. With more than a quarter (26.4%) TO of their primary HIV care settings and most (74.4%) of these coming from HCIII (36.6%) and hospital (37.8%) levels, this might point to capacity gaps in managing such cases at those lower levels. CALHIV are diagnosed with AHD and transferred out to higher facilities for better management, or those diagnosed at the hospital level are later referred to lower facilities after stabilizing. The high proportion transferred out and/or LTFU are often associated with poor documentation of treatment outcomes[17,18] and as such could have underestimated the mortality rate in our study.

A higher proportion (15.9%) who died among those on ART six months or less, was concerning and suggest existing health system gaps to capture this age group much earlier or to provide high quality of care after AHD diagnosis. This includes the abilities to screen and manage opportunistic infections like TB, cryptococcal meningitis, and other severe bacterial infections as well as ensuring adherence to prescribed medications.

CALHIV with moderate and severe acute malnutrition had 24.5% and 29.1% respective prevalences of AHD compared to those without malnutrition at 16.5%. Twenty-one percent of those with AHD and severe acute malnutrition died compared to 3% among those with no malnutrition. Our study could not elicit if malnutrition is a cause or an effect of AHD but these findings underscore the increased proportion of deaths if one had both AHD and severe malnutrition. Studies elsewhere reported similarly significant high rates of death (18-28%) among children with severe malnutrition as compared to those without[19,20]. The risk of death is even higher among severely malnourished children under five years old living with HIV.

Our study is one of the few, if not the only study, to have quantified the risk of mortality among CALHIV with AHD at 3.41 per 1,000 person-years. Results in Table 4 highlighting higher risk of mortality among CALHIV with malnutrition suggests the need to explore the effect of malnutrition on mortality rates among CALHIV diagnosed with AHD. The increased risk of death particularly among those who had been on ART for less than 6 months highlights the importance of the first year on ART in changing the tide and reducing AHD-related mortalities in this age group. Existing risk of death (Figure1) even after 24 months on ART is another finding that suggests that not following the national guidelines recommendations to improve ART adherence and continue screening and prophylaxis for opportunistic infections could have negative impacts on the health and lives of CALHIV.

**Table 4.**
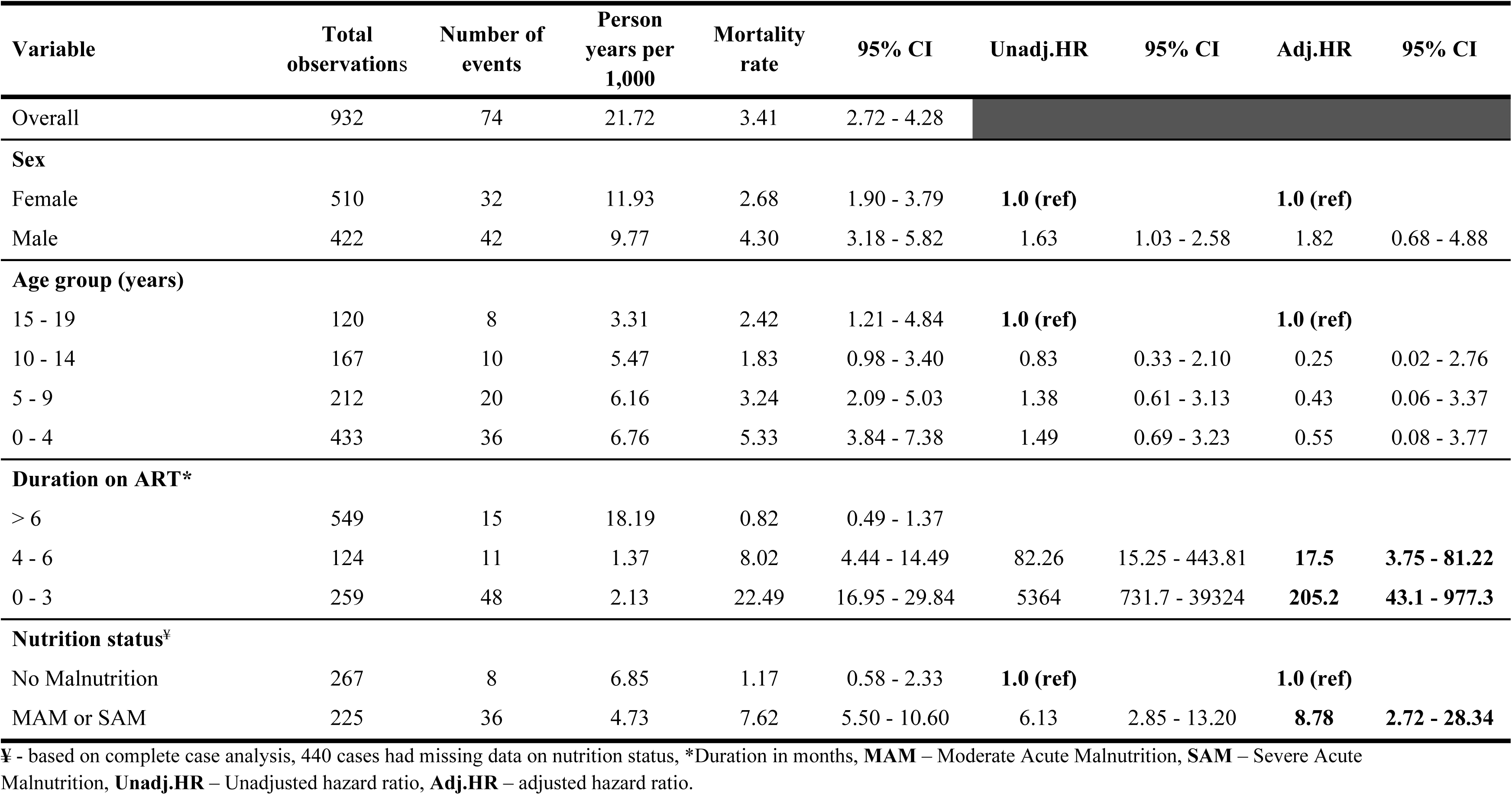
Mortality rate and associated factors of mortality among CALHIV diagnosed with Advanced HIV Disease between January 2016 - July 2023 in Uganda.

| Variable | Total observations | Number of events | Person years per 1,000 | Mortality rate | 95% CI | Unadj.HR | 95% CI | Adj.HR | 95% CI |
| --- | --- | --- | --- | --- | --- | --- | --- | --- | --- |
| Overall | 932 | 74 | 21.72 | 3.41 | 2.72 - 4.28 |  |  |  |  |
| **Sex** |  |  |  |  |  |  |  |  |  |
| Female | 510 | 32 | 11.93 | 2.68 | 1.90 - 3.79 | **1.0 (ref)** |  | **1.0 (ref)** |  |
| Male | 422 | 42 | 9.77 | 4.30 | 3.18 - 5.82 | 1.63 | 1.03 - 2.58 | 1.82 | 0.68 - 4.88 |
| **Age group (years)** |  |  |  |  |  |  |  |  |  |
| 15 - 19 | 120 | 8 | 3.31 | 2.42 | 1.21 - 4.84 | **1.0 (ref)** |  | **1.0 (ref)** |  |
| 10 - 14 | 167 | 10 | 5.47 | 1.83 | 0.98 - 3.40 | 0.83 | 0.33 - 2.10 | 0.25 | 0.02 - 2.76 |
| 5 - 9 | 212 | 20 | 6.16 | 3.24 | 2.09 - 5.03 | 1.38 | 0.61 - 3.13 | 0.43 | 0.06 - 3.37 |
| 0 - 4 | 433 | 36 | 6.76 | 5.33 | 3.84 - 7.38 | 1.49 | 0.69 - 3.23 | 0.55 | 0.08 - 3.77 |
| **Duration on ART\*** |  |  |  |  |  |  |  |  |  |
| > 6 | 549 | 15 | 18.19 | 0.82 | 0.49 - 1.37 |  |  |  |  |
| 4 - 6 | 124 | 11 | 1.37 | 8.02 | 4.44 - 14.49 | 82.26 | 15.25 - 443.81 | **17.5** | **3.75 - 81.22** |
| 0 - 3 | 259 | 48 | 2.13 | 22.49 | 16.95 - 29.84 | 5364 | 731.7 - 39324 | **205.2** | **43.1 - 977.3** |
| **Nutrition status<sup>‡</sup>** |  |  |  |  |  |  |  |  |  |
| No Malnutrition | 267 | 8 | 6.85 | 1.17 | 0.58 - 2.33 | **1.0 (ref)** |  | **1.0 (ref)** |  |
| MAM or SAM | 225 | 36 | 4.73 | 7.62 | 5.50 - 10.60 | 6.13 | 2.85 - 13.20 | **8.78** | **2.72 - 28.34** |

<sup>‡</sup> - based on complete case analysis, 440 cases had missing data on nutrition status, \*Duration in months, **MAM** – Moderate Acute Malnutrition, **SAM** – Severe Acute Malnutrition, **Unadj.HR** – Unadjusted hazard ratio, **Adj.HR** – adjusted hazard ratio.

### Study Limitations

Our study had some limitations. First, having a high proportion of missing viral load values especially among CALHIV under five years old (225, 36%) could have overestimated or underestimated the prevalence of AHD in this age group. However, given that the number of children under five years who were on ART over 12 months and did not have any viral load value was low (24), our estimates of AHD are likely to be accurate. We also used a cutoff of 1,000 copies/ml for viral suppression in this study which could have also overestimated the prevalence of AHD among those under five years old. There were no data on other opportunistic infections like cryptococcal meningitis and pneumonia that also contribute to mortality due to AHD[21].

### Conclusion

Results show prevalence of AHD among CALHIV to be 18.2% and pronounced among those under 5 years and those on ART ≤12 months. CALHIV with AHD and with malnutrition are more likely to die even when on ART. Further research is needed to determine whether malnutrition is a cause or an effect of AHD as well as the contribution of transfer out and/or LTFU on the true estimates of AHD and mortality among CALHIV.

## Supporting Information

**S1 file**. Uganda Ministry of Health 2022 ART guidelines

## Data Availability

All relevant data are within the manuscript and its Supporting Information files.

## Acknowledgements

We would like to thank all the monitoring and evaluation officers and health information system officers under the ACE-FORT and ACE-Hoima projects in mid-western Uganda for their efforts in ensuring timely and accurate update of the EMR in different health facilities. Additional thanks go to Dr Patricia Nahirya Ntege for her stewardship as the research directorate at Baylor-Uganda, and to the technical leads for each thematic area at CDC Uganda for their routine insightful guidance on project implementation to achieve project targets as well as improve the quality of lives of the project beneficiaries. In a special way, we would like to acknowledge Rachel Namuddu at Baylor-Uganda as well as Kenneth Mwambi at CDC-Uganda for spearheading the research administration and ethical clearances at all levels. The findings and conclusions in this report are those of the authors and do not necessarily represent the official position of the Centers for Disease Control and Prevention (CDC) or PEPFAR.

## Author contribution

**Conceptualization**: Calvin Epidu, Rogers N. Ssebunya

**Data curation**: Edgar Sserunkuma, Emmanuel Tumwine, Patrick Kizza

**Methodology and Formal analysis**: Rogers Nelson Ssebunya, Freddrick E. Makumbi

**Project administration**: Michael Juma, Betty Nsangi, Albert K. Maganda, Denise J. Birungi, Henry Balwa, Dithan Kiragga

**Writing original draft**: Rogers N. Ssebunya

**Review & editing**: Rogers N. Ssebunya, Calvin Epidu, Henry Balwa, Betty Nsangi, Arthur G. Fitzmaurice, Dithan Kiragga. All Authors read and approved the manuscript.

